# DNA Methylation Profiling in Childhood-Onset Lupus Reveals Distinct Epigenetic Clusters and Suggests Epigenetic Drivers of Disease Activity

**DOI:** 10.1101/2025.09.30.25337006

**Authors:** Desiré Casares-Marfil, Gülşah Kavrul Kayaalp, Vafa Guliyeva, Özlem Akgün, Şeyma Türkmen, Elif Kılıç Könte, Seher Şener, Sezgin Şahin, Özgür Kasapçopur, Betül Sözeri, Selçuk Sözer Tokdemir, Seza Özen, Nuray Aktay Ayaz, Amr H Sawalha

## Abstract

**Objectives:** Systemic lupus erythematosus (SLE) is a chronic autoimmune disease affecting multiple organs, with childhood-onset SLE (cSLE) typically presenting a more severe course and greater genetic risk than adult-onset SLE. While DNA methylation plays a key role in lupus pathogenesis, the epigenetic landscape of cSLE remains understudied. This study aimed to investigate DNA methylation changes in cSLE.

**Methods:** A total of 64 cSLE patients and 47 healthy control DNA samples isolated from peripheral blood mononuclear cells (PBMCs), along with an independent validation cohort of 38 patient DNA samples from whole blood, were analyzed. DNA methylation was assessed using the Infinium MethylationEPIC v2.0 array (Illumina), with quality control and statistical analyses performed using the *minfi* and *limma* R packages. Methylation differences were tested via linear regression adjusting for age, sex, medication use, and cell composition. Clinical features were compared using chi-square test or Fisher’s exact test, and gene ontology enrichment was conducted using Metascape and GREAT.

**Results:** Differential methylation analysis revealed significant hypomethylation in interferon-regulated genes (e.g., *DTX3L*, *PARP9*, *IFI44L*, *MX1*), enriched in type I interferon-related processes. Hypomethylation in genes linked to B cell activation and senescence correlated with higher SLEDAI scores. K-means clustering identified three distinct methylation-based cSLE subgroups, each enriched for different biological processes: cell adhesion/growth factor response (Cluster One), cell differentiation/fate (Cluster Two), and oxidative stress/Rap1 signaling (Cluster Three). Sex-based analysis showed immune-related hypomethylation in male patients, with over 80% of these sites validated in the replication cohort.

**Conclusion:** cSLE displays distinct DNA methylation patterns associated with disease activity, molecular subgroups, and sex, underscoring the potential for epigenetically informed diagnostics and therapies.

## INTRODUCTION

Systemic lupus erythematosus (SLE), or lupus, is an autoimmune disease that affects multiple organs and systems, potentially leading to significant organ damage, disability, and mortality. It is characterized by the generation of autoantibodies targeting various autoantigens, particularly nuclear components. The formation and deposition of immune complexes in different organs result in inflammation and subsequent organ damage. Despite advances in our understanding of lupus pathogenesis, the molecular mechanisms that underlie the distinct immunophenotype and clinical heterogeneity of the disease remain incompletely understood.

Genetic factors play a crucial role in the etiology of SLE, as evidenced by twin concordance rates, familial clustering, and the identification of more than 180 genetic risk loci for SLE ^1^. Similarly, epigenetic dysregulation plays an important role in the pathogenesis of SLE. We have previously demonstrated that hypomethylation is particularly enriched in interferon-related genes in patients with SLE ^2, 3^. Further, we demonstrated significant correlations between DNA methylation changes and disease activity in SLE ^4^. The association between SLE manifestations, such as skin involvement or nephritis, with specific DNA methylation changes suggests a potential role for epigenetic changes in explaining disease heterogeneity in SLE ^5, 6^.

Onset before the age of 18 is defined as childhood-onset SLE (cSLE), accounting for approximately 15-20% of all SLE cases ^7^. Childhood-onset SLE exhibits a more aggressive disease course with increased frequency of organ involvement compared to adult-onset disease^8^. Furthermore, childhood-onset SLE is characterized by a higher cumulative genetic risk compared to adult-onset disease ^8^. While DNA methylation changes in adult-onset SLE are well-documented, their contribution to cSLE pathogenesis requires further exploration.

In this study, we performed a comprehensive genome-wide DNA methylation analysis in a well-characterized cohort of patients with cSLE and matched healthy controls. We identified differentially methylated positions (DMPs) and regions (DMRs) associated with disease status, disease activity, and sex, and further characterized epigenetic subtypes of cSLE through unsupervised clustering. Our findings highlight robust hypomethylation at interferon-regulated loci, epigenetic correlates of disease activity, and distinct methylation-based patient clusters associated with clinical and immunological features. These data offer novel insights into the epigenetic architecture of cSLE and provide a framework for stratifying patients based on molecular epigenetic signatures.

## METHODS

### Study cohort and demographics

This study included two cohorts of cSLE patients and healthy controls from Turkey. The primary discovery cohort comprised a total of 64 patients and 47 age- (±5 years) and sex-matched healthy individuals, while the independent validation cohort included 38 patients. All patients included in this study fulfilled the 1997 revised American College of Rheumatology classification criteria for SLE ^9^ and had disease onset before 18 years of age. The study cohort patients were regularly followed at three pediatric rheumatology centers in Istanbul, while the validation cohort was recruited from a single pediatric center in Ankara. The recruiting centers receive referrals from across Western Turkey, and all patients and controls are of Turkish ancestry.

Clinical and laboratory data were retrospectively collected using a standardized form across all centers. Extracted information included demographic characteristics, including age at disease onset and at last visit, cumulative ACR classification criteria, other clinical and immunological findings observed during the disease course, and medications administered at the last follow-up visit. Disease activity was assessed using the Systemic Lupus Erythematosus Disease Activity Index 2000 revision (SLEDAI-2K), and organ damage was evaluated with the Systemic Lupus International Collaborating Clinics/American College of Rheumatology (SLICC/ACR) Damage Index (SDI). When available, renal biopsy findings were classified based on the 2003 International Society of Nephrology/Renal Pathology Society (ISN/RPS) criteria for lupus nephritis. Neurological involvement was defined based on the ACR nomenclature and case definitions for neuropsychiatric SLE ^10^. The last visit corresponds to the time point at which blood samples were collected for DNA isolation. Protocols used in the study followed the ethical guidelines of the 1975 Declaration of Helsinki, and all individuals included signed written informed consents. Ethics approval for this study was obtained from the Istanbul University

Ethics Committee (approval number: 2022/1480561), and the study was approved by the institutional review board of the University of Pittsburgh.

### Sample collection and DNA isolation

Peripheral blood (10 ml) was collected into EDTA-containing tubes from each participant. In the discovery cohort, peripheral blood mononuclear cells (PBMCs) were isolated on the same day using Ficoll-Paque density gradient centrifugation, followed by genomic DNA extraction with the Quick-DNA Miniprep Plus Kit (Zymo Research, USA) according to the manufacturer’s protocol. For the validation cohort, genomic DNA was directly extracted from whole blood samples collected in EDTA tubes. For all samples, DNA was eluted in water and quantified using Qubit Fluorometric Quantification (ThermoFisher).

### DNA methylation profiling

A total of 300 ng of DNA from each sample was bisulfite-converted using the EZ-96 DNA Methylation kit (Zymo Research) following manufacturer’s instructions. Converted DNA was hybridized to the Infinium MethylationEPIC v2.0 BeadChip array (Illumina, Inc., San Diego, CA, USA) to assess over 930,000 DNA methylation sites across the genome. Sample hybridization and array scanning were performed at the University of Michigan Advanced Genomics Core.

### Quality controls and normalization

DNA methylation raw data were processed using *minfi* R/Bioconductor package (v1.52.1) for quality control and normalization steps ^11^. Probes located in single-nucleotide polymorphisms (SNPs), CH probes (non-CpG methylation), those in sex chromosomes, and probes with a detection p-value > 0.01 were removed in each cohort separately. In addition, those samples with discrepancies between recorded sex information and estimated sex from the data were excluded from further analysis. The Quantile and Noob normalization methods were applied. Genomic build GRCh38/hg38 was used for probe annotation ^12^. Since DNA methylation patterns may vary among cell types, we estimated and adjusted subsequent statistical analyses for cell type composition using DNA methylation-based deconvolution. Cell-type proportions were estimated using reference-based deconvolution with the Houseman algorithm ^13^, as implemented in the *ewastools* and *meffil* packages ^14, 15^.

DNA methylation was quantified using both β values, which indicate the proportion of methylation on a scale from 0 to 1, and M values, which are calculated as the log2 transformation of the β values. While M values were employed for statistical testing, β values were used to aid interpretation and for data visualization. To evaluate for potential cofounding effects and correlation between covariates, principal component analysis (PCA) was conducted to calculate eigenvalues using patient medication data at last visit for prednisolone, azathioprine, hydroxychloroquine, mycophenolate mofetil, and rituximab, as well as for estimated cell composition, including monocytes, B cells, CD4^+^ T cells, CD8^+^ T cells, granulocytes, NK cells and nucleated red blood cells. Those principal components that comprise at least 90% of the cumulative explained variance were used as covariates in subsequent analyses.

### Clustering analysis of childhood-onset lupus patients

To characterize the distribution of cSLE patients based on their DNA methylation profiles, we performed a k-means clustering analysis of patients from the discovery cohort to identify groups of patients that may reflect underlying biological differences or molecular subtypes. Methylation β values were adjusted for age, sex, cell composition PCs, and medication PCs, using linear modeling to account for potential confounding factors. The adjusted matrix of β values was used as input for PC dimensional reduction of the data (**Supplementary Figure 1**) and unsupervised clustering using the k-means algorithm, implemented with the *kmeans* R function. The optimal number of clusters was determined by evaluating the elbow and silhouette width methods.

### Differential methylation analysis

Probe-wise linear regressions were performed to detect DMPs that show methylation differences between the groups tested, i.e. cSLE patients and healthy controls, disease activity as measured by SLEDAI scores, and male and female patients, using the *limma* (v3.62.2) package ^16^. Age, sex, and the top 5 cell composition PCs were used as covariates in the case-control analysis, while the top 4 medication PCs were also included as covariates in case-case analyses. An empirical Bayes moderated t-statistic and P-value were calculated for each probe. Differential methylation (Δβ) between the groups in each comparison was estimated as the difference in mean β values for each CpG. Thus, CpG sites were considered significantly differentially methylated if they had a Benjamini-Hochberg FDR-adjusted p-value < 0.05 and were differentially methylated by at least 10% (|Δβ| > 0.1) between groups if the variable of interest was categorical. This threshold helps ensure the capture of biologically meaningful and consistent signal variation while minimizing false positives from technical noise or low-impact variability ^17^. In addition to linear regression, Pearson’s correlation test was used in the analysis of the relationship between SLEDAI scores and DNA methylation levels. To assess differences in DNA methylation patterns associated with each patient cluster, individuals in each cluster were compared against the combined set of individuals from the other two clusters: Cluster One vs clusters Two and Three; Cluster Two vs clusters One and Three; and Cluster Three vs clusters One and Two. These analyses were adjusted for sex, age, and principal components for cell composition and medications. In addition, differences in clinical manifestations between clusters were assessed using Fisher’s exact test or the chi-square test, depending on whether the number of individuals per comparison group was less than 5 or not, respectively. Finally, the presence of DMR was also assessed by using *DMRcate* ^18^. Those DMRs with a Stouffer adjusted p-value < 0.05 and containing at least 5 CpGs were considered significant.

### Enrichment analysis in biological process

Genes annotated to each significant CpG site were used to perform Gene Ontology (GO) and KEGG Pathway enrichment analyses through the Metascape web-based portal ^19^, in order to explore the potential biological processes associated with the genomic regions identified in our methylation analyses. Only those terms with a p-value < 0.01 and at least three genes were reported. An additional GO analysis was performed using the genomic positions of significantly methylated positions using the web-based portal GREAT ^20^.

## RESULTS

The median age among patients in the discovery cohort at disease onset was 12.17 years (interquartile range [IQR]: 9.77–14.38), while the median age at the last visit was 17.44 years (IQR: 15.17–18.60). The majority were female (n = 48, 75.0%), with a median disease duration of 3.97 years (IQR: 2.08–5.86). The most frequently observed cumulative clinical features were hematologic involvement (n = 42, 65.6%), malar rash (n = 41, 64.1%), arthritis (n = 35, 54.7%), and renal involvement (n = 31, 48.4%). At the time of the last clinical assessment, the most administered treatments included hydroxychloroquine (n = 55, 85.9%), mycophenolate mofetil (n = 26, 40.6%), and corticosteroids (n = 20, 31.3%). **Supplementary table 1** summarizes the demographic, clinical, immunological, and treatment-related characteristics of the study cohorts.

### DNA methylation differences in childhood-onset lupus

After quality control, a total of 880,024 CpGs were included for analysis. The analysis of DNA methylation differences between cSLE cases (n = 64) and healthy controls (n = 47) in the discovery cohort revealed a total of 20 DMPs (FDR-Adjusted p-value < 0.05 and |Δβ| > 0.1). All these positions were hypomethylated in cases and were annotated to 12 different genes (**Table 1**, **Figure 1**). These DMPs were predominantly located in promoter regions 200-1500 base pairs upstream of the transcription start site and in intronic regions. The majority of these DMPs were annotated to genes related to the type I interferon (IFN) signature, including *DTX3L*, *IFI44L*, *MX1*, *IFIT1*, *PLSCR1*, *DDX60*, *USP18*, *PARP9*, and *IRF9*. Gene Ontology (GO) and KEGG pathway enrichment analyses demonstrated significant overrepresentation of the type I interferon signaling pathway, antiviral response, and innate immune response-related pathways (**Figure 1, Supplementary Table 2**).

**Figure 1.**
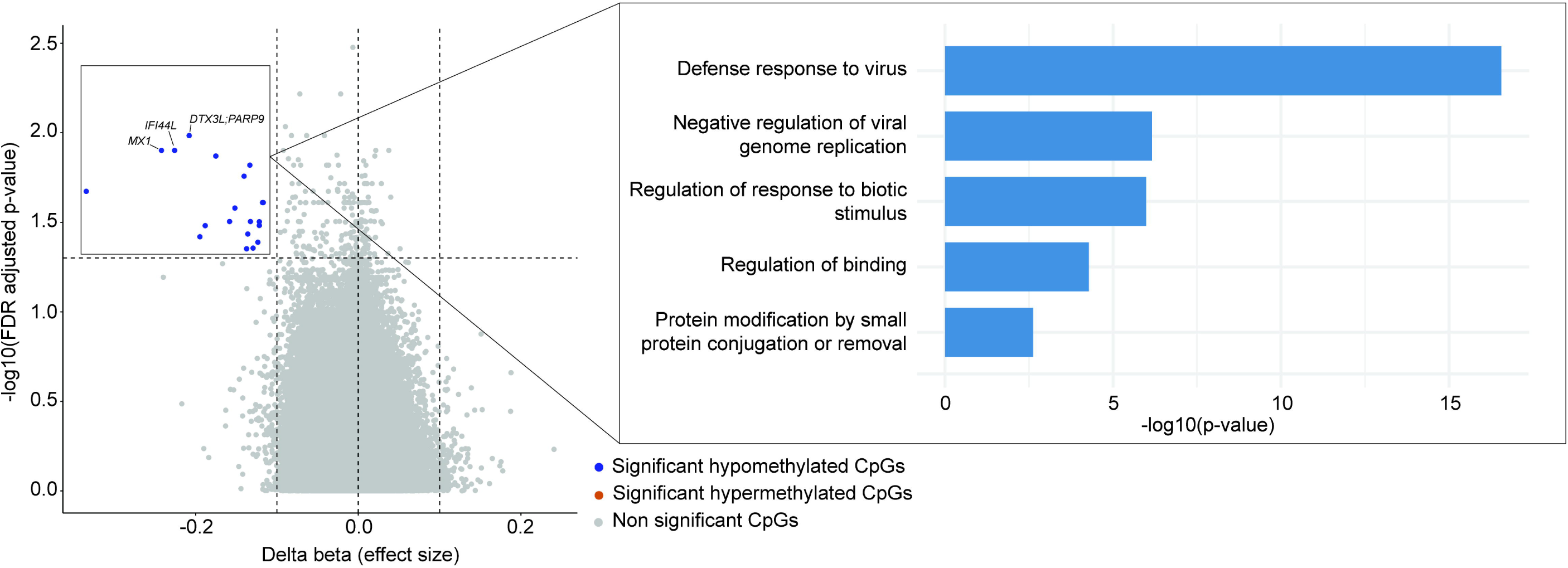
Volcano plot of differentially methylated positions (DMPs) between childhood-onset lupus patients (cSLE) and matched healthy controls in the discovery cohort. Significant DMPs that show hypomethylation in cSLE patients (blue) are differentially methylated by at least 10% between groups and with an FDR-adjusted p-value < 0.05. The three most significant hypomethylated DMPs were annotated to their nearest genes and labeled in the plot. No significant differentially methylated positions showing hypermethylation in cSLE cases (orange) were identified. On the right, gene ontologies (Biological Process) and Kyoto Encyclopedia of Genes and Genomes (KEGG) pathways main clusters enriched in the genes annotated to the hypomethylated significant differentially methylated positions are represented. Only terms with a p-value < 1x10^−3^ and at least three genes in the term were reported.

**Table 1.**
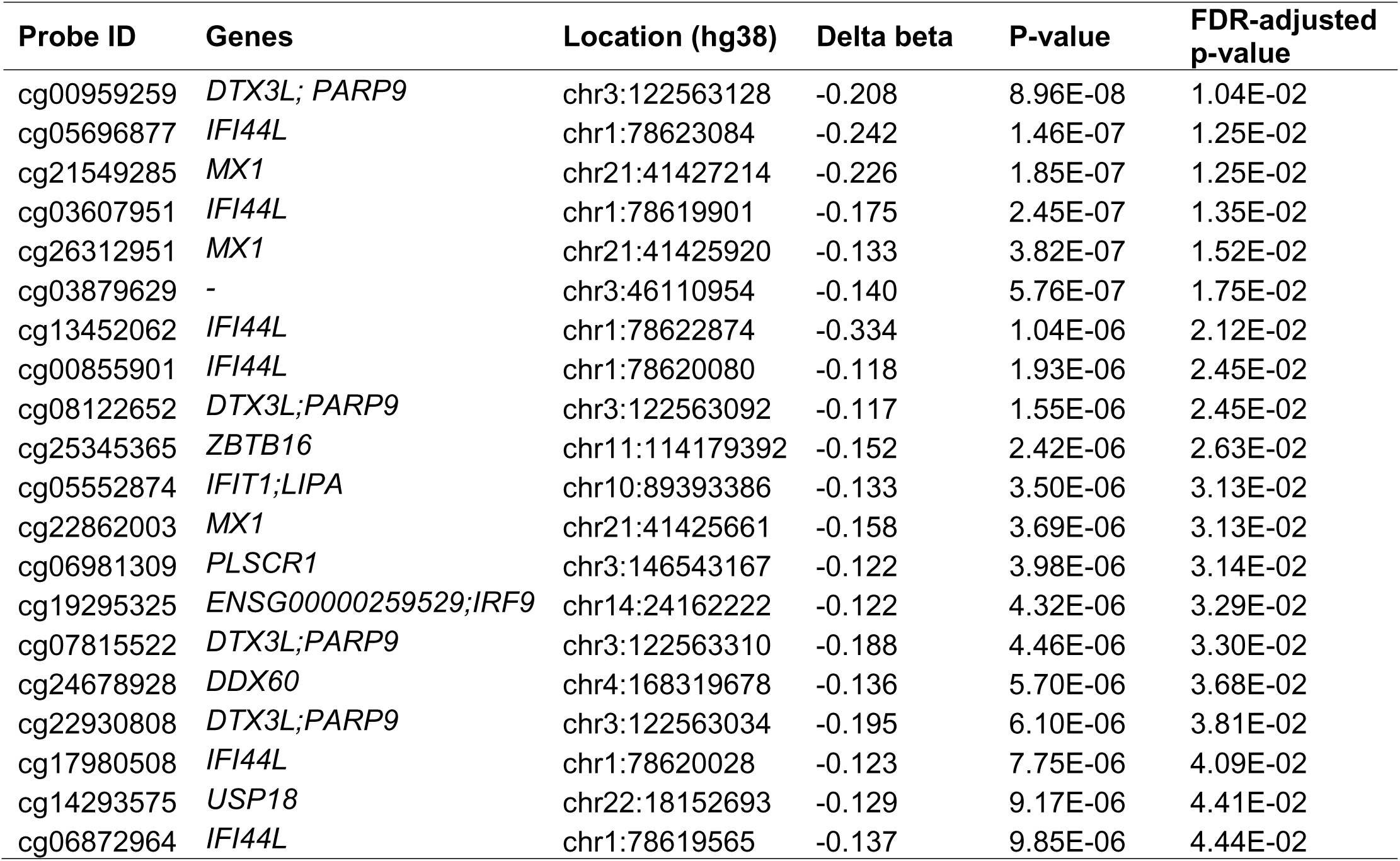
Differentially methylated positions (DMPs) between childhood-onset lupus patients and healthy.

| Probe ID | Genes | Location (hg38) | Delta beta | P-value | FDR-adjusted p-value |
| --- | --- | --- | --- | --- | --- |
| cg00959259 | *DTX3L; PARP9* | chr3:122563128 | -0.208 | 8.96E-08 | 1.04E-02 |
| cg05696877 | *IFI44L* | chr1:78623084 | -0.242 | 1.46E-07 | 1.25E-02 |
| cg21549285 | *MX1* | chr21:41427214 | -0.226 | 1.85E-07 | 1.25E-02 |
| cg03607951 | *IFI44L* | chr1:78619901 | -0.175 | 2.45E-07 | 1.35E-02 |
| cg26312951 | *MX1* | chr21:41425920 | -0.133 | 3.82E-07 | 1.52E-02 |
| cg03879629 | - | chr3:46110954 | -0.140 | 5.76E-07 | 1.75E-02 |
| cg13452062 | *IFI44L* | chr1:78622874 | -0.334 | 1.04E-06 | 2.12E-02 |
| cg00855901 | *IFI44L* | chr1:78620080 | -0.118 | 1.93E-06 | 2.45E-02 |
| cg08122652 | *DTX3L;PARP9* | chr3:122563092 | -0.117 | 1.55E-06 | 2.45E-02 |
| cg25345365 | *ZBTB16* | chr11:114179392 | -0.152 | 2.42E-06 | 2.63E-02 |
| cg05552874 | *IFIT1;LIPA* | chr10:89393386 | -0.133 | 3.50E-06 | 3.13E-02 |
| cg22862003 | *MX1* | chr21:41425661 | -0.158 | 3.69E-06 | 3.13E-02 |
| cg06981309 | *PLSCR1* | chr3:146543167 | -0.122 | 3.98E-06 | 3.14E-02 |
| cg19295325 | *ENSG00000259529;IRF9* | chr14:24162222 | -0.122 | 4.32E-06 | 3.29E-02 |
| cg07815522 | *DTX3L;PARP9* | chr3:122563310 | -0.188 | 4.46E-06 | 3.30E-02 |
| cg24678928 | *DDX60* | chr4:168319678 | -0.136 | 5.70E-06 | 3.68E-02 |
| cg22930808 | *DTX3L;PARP9* | chr3:122563034 | -0.195 | 6.10E-06 | 3.81E-02 |
| cg17980508 | *IFI44L* | chr1:78620028 | -0.123 | 7.75E-06 | 4.09E-02 |
| cg14293575 | *USP18* | chr22:18152693 | -0.129 | 9.17E-06 | 4.41E-02 |
| cg06872964 | *IFI44L* | chr1:78619565 | -0.137 | 9.85E-06 | 4.44E-02 |

To assess consistent methylation changes across the genome we performed a DMR analysis. A total of 295 significant regions with at least 5 CpGs and FDR-Adjusted p-value < 0.05 were identified when comparing cases and controls (**Supplementary Table 3**). Two hundred and seventy-two of these regions were hypomethylated in cases compared with controls, and 23 were hypermethylated, with a mean size of 8.25 CpGs per region. These regions were annotated to 247 genes, including some genes related to lupus pathogenesis, such as *IFN-AS1* and *ZBED9*, among the most significant regions.

### DNA methylation analysis across disease activity levels

We performed a differential methylation analysis to investigate the relationship between DNA methylation and disease activity, as measured by SLEDAI scores. After adjusting for age, sex, estimated cell proportion PCs, and medication PCs, we identified a total of 77 DMPs in the discovery cohort whose methylation levels were significantly associated with SLEDAI scores (FDR-Adjusted p-value < 0.05, **Figure 2**, **Supplementary Table 4**). The enrichment analysis of biological processes of the genes associated with significantly hypomethylated DMPs highlighted a correlation between B cell activation and cellular senescence with disease activity (**Figure 2**, **Supplementary Table 5**).

**Figure 2.**
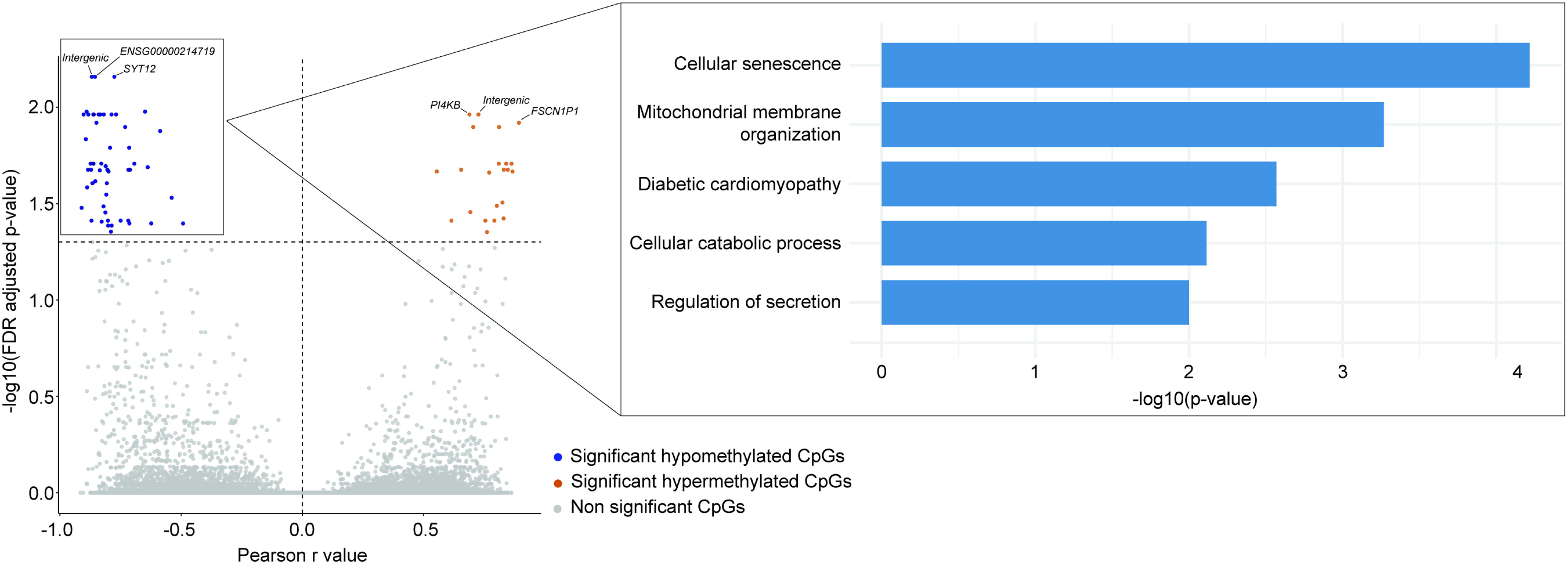
Volcano plot showing the correlation between DNA methylation levels and disease activity, as measured by the SLEDAI score, in childhood-onset lupus (cSLE) patients from the discovery cohort. Significant differentially methylated positions (DMPs) with an FDR-adjusted p-value < 0.05 are colored according to directionality, where blue indicates hypomethylation and orange indicates hypermethylation in association with increased disease activity. FDR-adjusted p-values were calculated using lineal regression. The three most significant hypo- and hypermethylated DMPs were annotated to their nearest genes and labeled in the plot. On the right, gene ontologies (Biological Process) and Kyoto Encyclopedia of Genes and Genomes (KEGG) pathways main clusters enriched in the genes annotated to the hypomethylated significant differentially methylated positions are represented. Only terms with a p-value < 1x10^−3^ and at least three genes in the term were reported.

### Sex-based comparison of DNA methylation in childhood-onset lupus

We assessed the influence of DNA methylation in disease differences between male and female cSLE patients. After adjusting for age, cell proportion PCs, and medication PCs, we identified 67 significant DMPs (FDR-Adjusted p-value < 0.05 and |Δβ| > 0.1) in the discovery cohort (**Figure 3**, **Supplementary Table 6**). Of these, 44 CpGs showed hypomethylation and 23 hypermethylation in male patients, located within 51 and 24 unique genes, respectively. Sex-based analysis in the validation cohort replicated 88% of the findings observed in the discovery cohort (**Supplementary Table 6**). Functional enrichment analysis of genes annotated to significantly hypomethylated DMPs revealed overrepresentation of pathways related to the regulation of the immune system (**Figure 3**, **Supplementary Table 7**).

**Figure 3.**
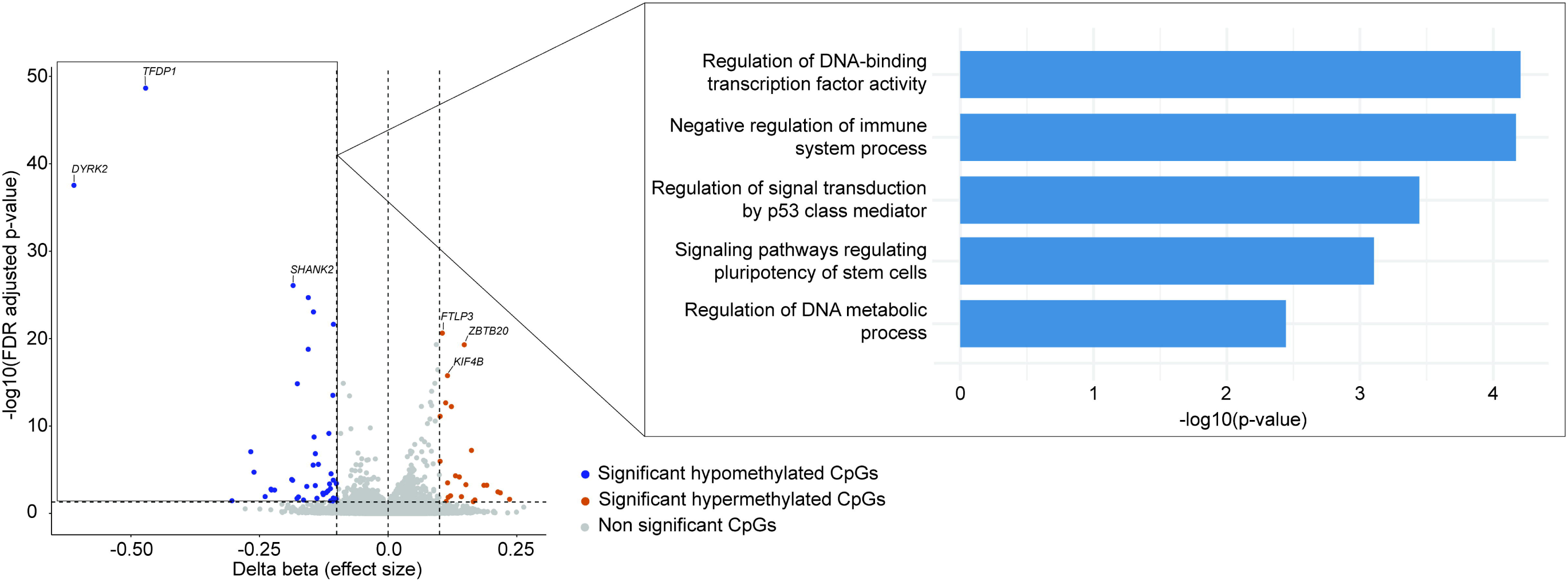
Volcano plot of differentially methylated positions (DMPs) between male and female childhood-onset lupus (cSLE) patients from the discovery cohort. Significant DMPs are differentially methylated by at least 10% between groups and with an FDR-adjusted p-value < 0.05. DMPs are colored according to directionality, where blue indicates hypomethylation and orange indicates hypermethylation in male cSLE patients. The three most significant hypo- and hypermethylated DMPs were annotated to their nearest genes and labeled in the plot. On the right, gene ontologies (Biological Process) and Kyoto Encyclopedia of Genes and Genomes (KEGG) pathways main clusters enriched in the genes annotated to the hypomethylated significant differentially methylated positions are represented. Only terms with a p-value < 1x10^−3^ and at least three genes in the term were reported.

We next identified sex-based DMRs. This approach uncovered a total of six significant DMRs (FDR-Adjusted p-value < 0.05 and at least 5 CpGs), annotated to four unique genes (**Supplementary Table 8**). The most significant DMR mapped to the *IFNG-AS1* gene, which encodes a long non-coding RNA that plays a role in regulating IFN-γ.

### Identification and characterization of DNA methylation clusters in lupus patients

A clustering analysis based on DNA methylation profiles from samples from the discovery cohort revealed three clusters of patients with different clinical and serological features (**Figure 4A, Supplementary Figure 2**). Cluster One comprised a total of 22 cSLE patients with a higher frequency of alopecia and leukopenia, and a lower rate of anti-RNP positivity compared to other clusters (**Supplementary Table 9**). This cluster was represented by 684 significant DMPs (FDR-Adjusted p-value < 0.05 and |Δβ| > 0.1), 180 hypomethylated and 504 hypermethylated (**Figure 4B**, **Supplementary Table 10**). These positions mapped to 157 and 469 unique genes for hypomethylated and hypermethylated positions, respectively. This cluster was characterized by enrichment in biological pathways related to cell adhesion and response to growth factor in hypomethylated genes (**Figure 5, Supplementary Table 11**). The second cluster identified consisted of 25 patients and exhibited higher renal involvement and anti-RNP positivity, but fewer constitutional symptoms, photosensitivity, hematologic manifestations, and leukopenia (**Supplementary Table 9**). The final visit SLEDAI-2K scores were higher in patients belonging to Cluster Two than in those from the other clusters. The analysis of methylation patterns defining this cluster revealed 18 hypomethylated and 33 hypermethylated positions differentially methylated in this cluster (**Figure 4B**, **Supplementary Table 10**). These positions mapped to 23 and 34 genes for hypomethylated and hypermethylated positions, respectively. The enrichment analysis of genes linked to hypomethylated positions revealed regulation of cell differentiation and cell fate determination pathways (**Figure 5, Supplementary Table 11**). Finally, Cluster Three consisted of 18 patients. Patients in this cluster did not show significant differences in clinical or immunological features compared to the other clusters (**Supplementary Table 9**). The analysis of methylation patterns representing this cluster revealed a total of 624 significant hypomethylated and 122 hypermethylated DMPs (**Figure 4B, Supplementary Table 10**), mapping to 113 and 577 unique genes, respectively. This cluster was characterized by an enrichment in response to oxidative stress and Rap1 signaling pathway in hypomethylated genes (**Figure 5, Supplementary Table 11**). The majority of DMPs identified in the three clusters were located in CpG islands and the regions 1500 bp upstream to the transcription start site of their nearest genes (**Supplementary Figure 3**).

**Figure 4.**
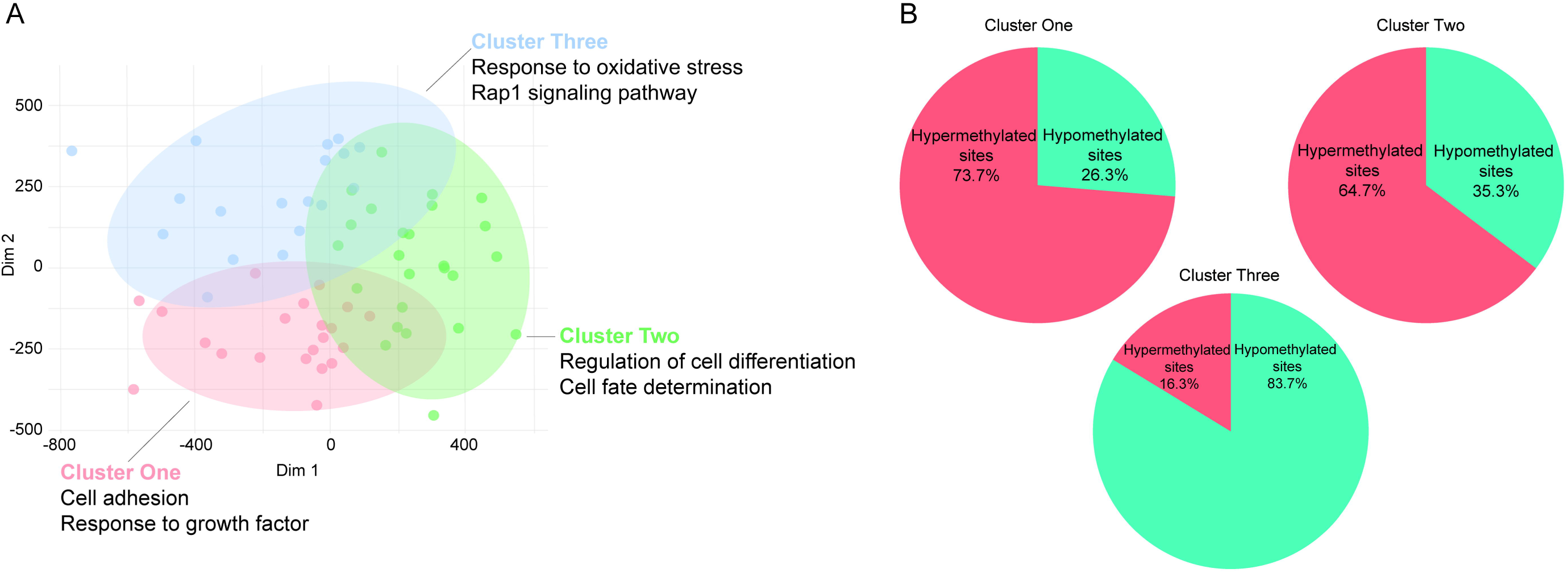
Childhood-onset lupus (cSLE) patient stratification based on DNA methylation patterns. (A) Scatter plot showing the distribution of cSLE patients from the discovery cohort according to k-means clustering method based on their DNA methylation profiles. Patients were grouped into three clusters, i.e. Cluster One (red), Cluster Two (green), and Cluster Three (blue). (B) Pie charts showing the percentage of hypermethylated (red) and hypomethylated (blue) significant differentially methylated positions (DMPs) on each DNA methylation cluster.

**Figure 5.**
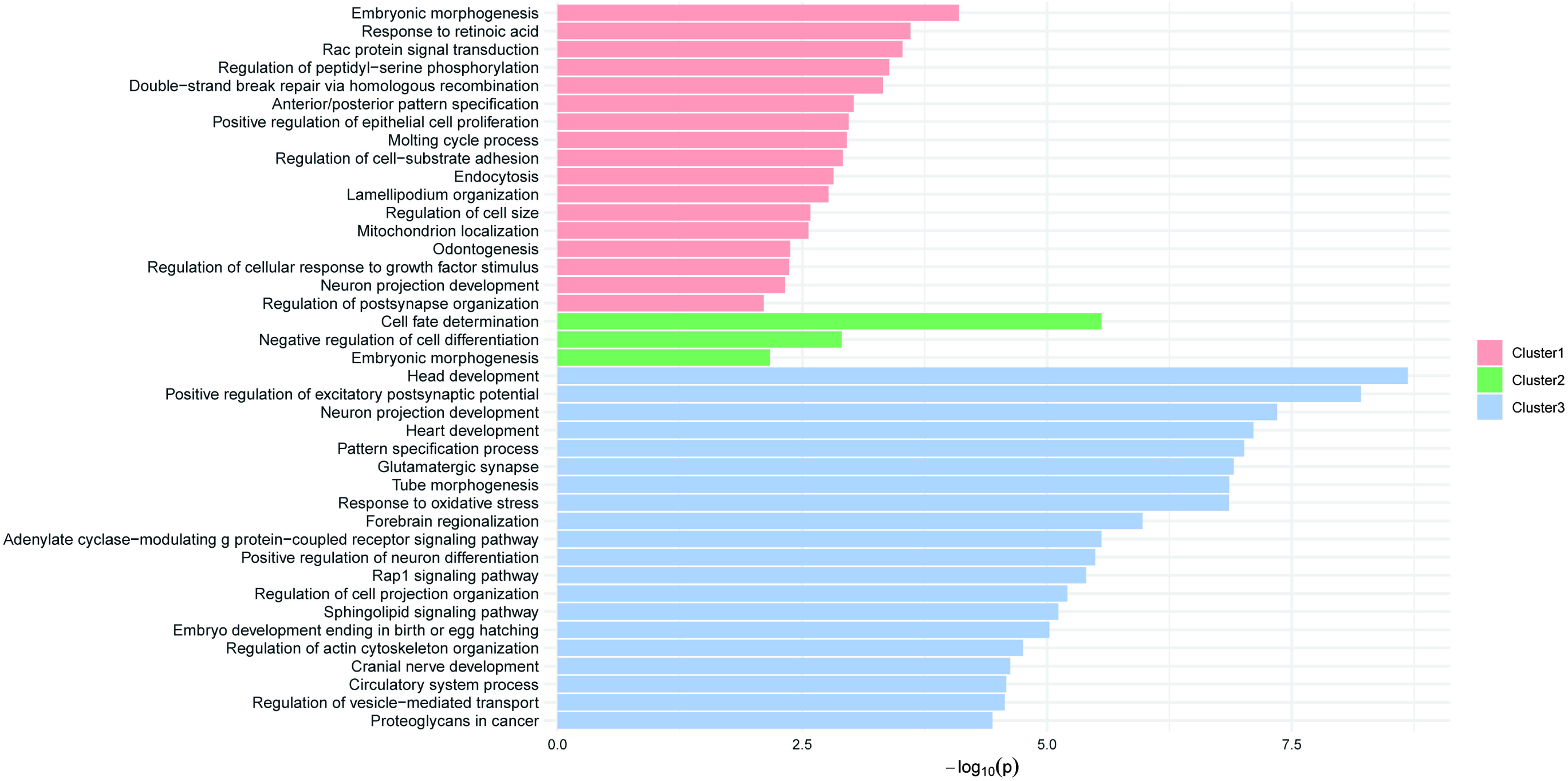
Gene ontologies (Biological Process) and Kyoto Encyclopedia of Genes and Genomes (KEGG) pathways identified from significantly hypomethylated differentially methylated positions (DMPs) within each DNA methylation cluster of childhood-onset lupus (cSLE) patients (Cluster One-red, Cluster Two-green, Cluster Three-blue). Only terms with a p-value < 1x10^−3^ and at least three associated genes are shown.

## DISCUSSION

We conducted a genome-wide comparison of DNA methylation profiles between a large, clinically well-characterized cohort of cSLE patients and age- and sex-matched healthy controls. While a previous study analyzed a small cohort of pediatric-onset SLE patients using purified immune cell lineages and whole blood, and identified differential DNA methylation across immune cell types ^21^, our work expands on these findings by integrating a broader analysis of epigenetic patterns in relation to clinical heterogeneity. Our findings underscore the potential utility of epigenetic profiling to refine molecular classification of lupus and guide precision medicine approaches in this disease.

We observed marked hypomethylation of IFN-stimulated genes in cSLE, consistent with previous reports from adult SLE studies. *IFI44L*, emerged as one of the most significantly hypomethylated genes and has previously been proposed as a promising epigenetic biomarker for SLE ^22^. Other IFN-regulated genes—including *MX1*, *DTX3L*, *IFIT1*, and *PARP9*—also showed robust hypomethylation, in line with prior epigenome-wide association studies ^23^. These results underscore that IFN-related epigenetic changes, previously characterized in adult-onset lupus, are likewise a prominent and defining feature of cSLE.

These results are consistent with findings reported in other type I IFN-opathies such as Aicardi-Goutières Syndrome (AGS), a rare monogenic disorder characterized by neurological involvement and shares clinical and molecular features with SLE. Similar to our findings in cSLE, a study in AGS reported robust demethylation in interferon-regulated genes, such as *IFI44L* ^24^. While the IFN methylation signature is well established in SLE, comparing methylation patterns with a monogenic disease like AGS may aid in refining molecular signature selection to better predict organ involvement in a more common and heterogeneous condition such as SLE. Unfortunately, our study was underpowered to detect significant DNA methylation changes in a case-case subset analysis focused on cSLE with neurological involvement (data not shown).

Beyond the IFN axis, we identified additional hypomethylated genes with immunoregulatory functions. *ZBTB16*, encoding the transcriptional regulator PLZF, emerged as a notable non-IFN gene. PLZF regulates lymphocyte development and modulates gene expression through interactions with histone deacetylases HDAC1 and HDAC3, with implications for immune homeostasis ^25–28^. Another significant non-IFN gene, *LIPA*, encodes lysosomal acid lipase, whose activity supports anti-inflammatory signaling via the generation of endogenous ligands that activate LXR and PPARδ pathways ^29^. The observed hypomethylation in cSLE in this gene may reflect a transcriptional response to increased apoptotic cell burden, although functional validation is needed.

DNA methylation analysis in relation to disease activity identified several genes previously implicated in SLE pathogenesis, including *PLA2R1* and *ANKS1A*, which exhibited progressive hypomethylation with increasing disease activity. *PLA2R1* encodes the phospholipase A2 receptor 1, a protein implicated in kidney disease. Genetic variants in this gene have previously been associated with SLE, specifically with lupus nephritis ^30^. Similarly, genetic polymorphisms within the *ANKS1A* gene, which encodes the ankyrin repeat and sterile alpha motif domain containing 1A, have been associated with serum IFN-α activity in SLE patients ^31^. Interestingly, despite their marked hypomethylation in cSLE patients, IFN-related genes did not show a correlation with disease activity. This is consistent with previous reports, suggesting that DNA methylation status of IFN-related genes remains relatively stable across different disease states^3^. In addition, the correlation between SLEDAI scores and hypomethylated genes could not be replicated in the validation cohort. This discrepancy may be attributable to differences in disease activity between the cohorts, as the majority of patients in the validation cohort had clinically inactive disease. Further, DNA methylation was assessed in peripheral blood mononuclear cells in the discovery cohort and in whole blood in the validation cohort.

Importantly, genes that progressively demethylate with higher SLEDAI scores were enriched in pathways related to cellular senescence, mitochondrial membrane organization, and B-cell activation. Cellular senescence is associated with the abnormal production of cytokines, growth factors, and proteases ^32^. A recent study further supports this concept by identifying senescence-related genes associated with lupus nephritis ^33^. Our analysis revealed hypomethylation of multiple genes implicated in senescence and B-cell activation—such as *NBN*, *MYH9*, *RBL1*, and *HDAC4*—in patients with higher disease activity, highlighting the involvement of these biological processes in active cSLE. The hypomethylation of *NBN*, a key component of the MRN complex involved in double-strand DNA break repair, points to persistent genotoxic stress and activation of DNA damage response mechanisms in patients with active disease ^34^. *MYH9*, which plays a role in cytoskeletal integrity and cell morphology, has also been previously associated with lupus nephritis susceptibility ^35^. RBL1 (p107), a member of the retinoblastoma family, acts as a key regulator of cell cycle checkpoints and senescence ^36^.

*HDAC4*, a histone deacetylase, modifies chromatin structure to regulate transcription factor access and is involved in glucose metabolism, cellular senescence, cell apoptosis, autophagy and inflammatory response ^37^. Collectively, these findings indicate that cellular senescence and B-cell activation through senescence-associated pathways contribute substantially to disease activity in cSLE. Genes associated with mitochondrial membrane organization (e.g. *SLC25A4* or *STPG1*) also exhibited hypomethylation correlated with disease activity. Mitochondria play a central role in maintaining cellular homeostasis, and their dysfunction has been implicated in increased reactive oxygen species (ROS) production and activation of the innate immune system in SLE ^38, 39^. Mitochondrial nucleic acids are highly antigenic, and their release into the cytosol—triggered by oxidative stress, environmental factors, or impaired autophagy—can have potent interferogenic effects ^40^. Disruptions in mitochondrial membrane organization may disturb this balance, potentially exacerbating disease activity in SLE ^41^.

Clustering based on DNA methylation profiles revealed three epigenetically distinct subgroups of cSLE patients, reflecting the underlying epigenetic heterogeneity of cSLE. In Cluster 1, hypomethylation was particularly notable in *KRAS*, *NF1*, and *AXL*—mediators of Ras/Rac1 pathways involved in cytoskeletal remodeling and immune regulation ^42^. *KRAS* mutations are known to cause Noonan-like RASopathies that can present with lupus-like autoimmunity ^43^. In addition, genes such as *RPTOR* and *TFRC*, involved in mTOR signaling and cellular metabolism, also showed hypomethylation in this cluster. Cluster Two was defined by hypomethylation in genes enriched in differentiation and cell fate determination pathways.

Notably, several of these genes—including *FOXG1*, *ISL1*, and *PRRX1*—have been shown to interact with or be regulated by the Wnt/β-catenin signaling pathway, which is involved in the pathogenesis of lupus ^44^. Cluster Three was enriched in pathways related to oxidative stress and Rap1 signaling, both of which intersect with MAPK and ERK signaling cascades. Notably, this cluster included genes associated with MAPK signaling and regulation of ERK1 and ERK2. Inhibition of ERK signaling has been shown to enhance the expression of methylation-sensitive genes implicated in lupus, suggesting a potential epigenetic mechanism linking ERK modulation to autoimmunity ^45^. Furthermore, oxidative stress—also enriched in this cluster—is known to affect ERK/MAPK signaling ^45^.

The analysis of sex-biased DNA methylation differences in cSLE revealed several hypomethylated genes in male patients compared to female patients, with a substantial proportion of these differences also replicated in an independent whole blood cohort. Sixteen DMPs that showed significant sex-associated differences in patients with cSLE also exhibited consistent sex differences in CD4⁺ T cells from adult-onset SLE patients ^46^. In our study, several genes related to transcription factors—most notably *TFDP1*, *HAND1*, *MYC*, and *TRIM24*—were found to be hypomethylated in cSLE male patients. In addition, some hypomethylated genes identified in our work have been previously associated with SLE. *MYC* encodes the transcription factor c-MYC, a key regulator of cell proliferation, metabolism, and apoptosis ^47^. Increased *MYC* expression has been reported in multiple tissues of lupus patients ^48, 49^, and its upregulation via the MAPK/ERK/p38 pathway was shown to promote glycolysis and drive the expansion of IL- 10⁺ regulatory B cells with proinflammatory features, ultimately contributing to pathogenic CD4⁺ T cell responses ^50^. *CARD8*, another hypomethylated gene in male patients, encodes a protein that modulates inflammasome assembly, pyroptosis, and IL-1β production ^51, 52^. Notably, a polymorphism in *CARD8* has been associated with lupus susceptibility specifically in male but not in female patients ^53^. Altogether, the coordinated hypomethylation and potential overactivation of these genes may reflect a cellular stress and related inflammation.

Our study has certain limitations. First, differences in cell composition between the discovery and validation cohorts may affect the comparability of results. While the validation cohort provided independent replication, the presence of neutrophils in whole blood samples could have influenced the validation of some findings. Replication in an independent PBMC cohort would more directly address this concern. We attempted to account for cellular heterogeneity in our samples by applying a deconvolution approach that infers relative cell-type composition from methylation signatures. However, deconvolution protocols have limitations and were not developed using pediatric age groups included in our study. Therefore, residual confounding by cell composition may still be present. Further, while whole blood samples may contain some cell-free DNA in proportions that are generally expected to be negligible, we cannot rule out that cell-free DNA affected our results in the validation cohort. Another limitation in our study is sample size. Although relatively large for genome-wide methylation studies, and in SLE and cSLE in particular, it remains limited for stratified sub-analyses by clinical manifestations.

Replication in larger cohorts is thus needed to robustly identify clinically relevant findings. Finally, while DNA methylation changes can influence gene expression, the absence of expression data in our study restricts functional characterization of the epigenetic differences observed.

In summary, we performed a comprehensive genome-wide DNA methylation analysis in cSLE. Our findings revealed distinct epigenetically defined patient clusters and methylation signatures associated with sex and disease activity, in addition to the interferon-related methylation patterns previously observed in adult SLE. By uncovering DNA methylation signatures linked to clinical heterogeneity, our results provide novel insight into the epigenetic landscape of cSLE and lay the groundwork for future studies aiming to identify epigenetic biomarkers and mechanistic pathways involved in disease pathogenesis.

## Data Availability

All data produced in the present work are contained in the manuscript

## ACKNOWLEDGEMENTS

This work was supported by the National Institute of Allergy and Infectious Diseases (NIAID) of the National Institutes of Health (NIH) grant number R01 AI097134. This research was supported in part by the University of Pittsburgh Center for Research Computing and Data, RRID:SCR_022735, through the resources provided. Specifically, this work used the HTC cluster, which is supported by NIH award number S10OD028483.

## AUTHOR CONTRIBUTIONS

Desiré Casares-Marfil: Writing – original draft, Formal analysis, Data curation. Gülşah Kavrul Kayaalp: Writing – original draft, Formal analysis, Data curation. Vafa Guliyeva: Data curation. Özlem Akgün: Data curation. Şeyma Türkmen: Data curation. Elif Kılıç Könte: Data curation. Seher Şener: Data curation. Sezgin Şahin: Data curation. Özgür Kasapçopur: Data curation. Betül Sözeri: Data curation. Selçuk Sözer Tokdemir: Data curation. Seza Özen: Data curation. Nuray Aktay Ayaz: Supervision, Resources, Project administration, Conceptualization, Data curation. Amr H Sawalha: Writing – review & editing, Supervision, Resources, Methodology, Funding acquisition, Conceptualization.

