## Supplementary Figures for "DNA Methylation Profiling in Childhood-Onset Lupus Reveals Distinct Epigenetic Clusters and Suggests Epigenetic Drivers of Disease Activity"

**Supplementary Figure 1.** Scatter plot showing the proportion of variance explained by each principal component (PC1–PC52). While 100% of the variance in the dimensionally-reduced data is explained by the first 52 PCs, at least 90% of this variance is captured by the first 43 PCs, which were used for the clustering analysis. The red line represents the cumulative variance explained, while the blue bars indicate the variance explained by individual components.

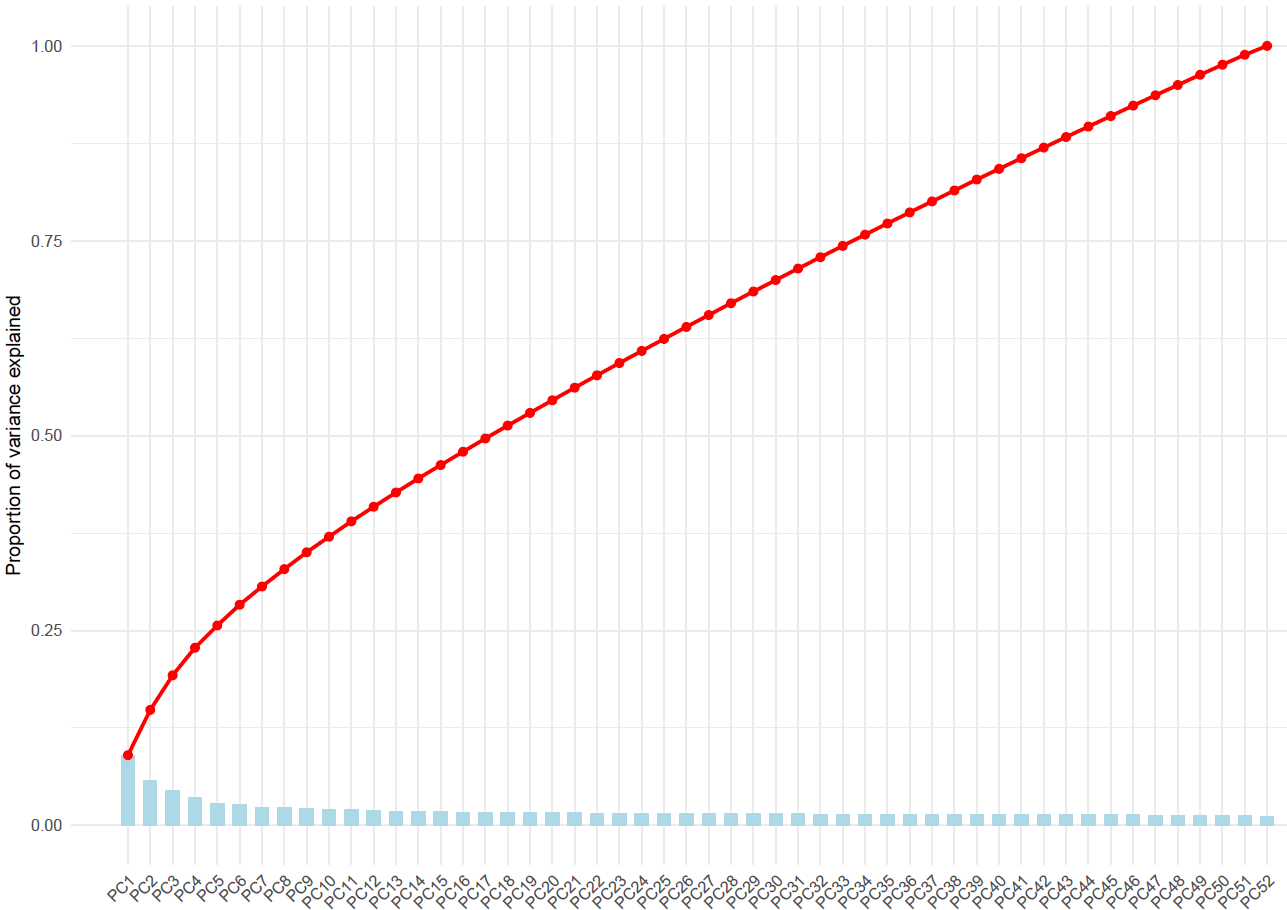

**Supplementary figure 2.** Silhouette plot showing the average silhouette scores for different numbers of clusters. The x-axis represents the number of clusters tested, and the y-axis represents the average silhouette width. The highest peak is observed at 3 clusters, indicating the optimal clustering configuration.

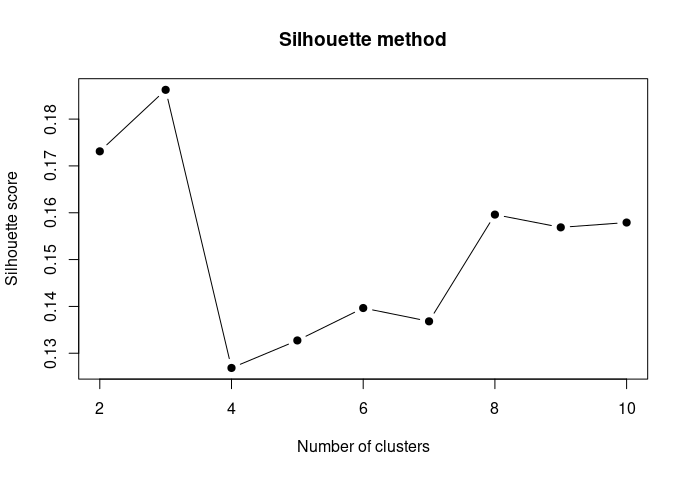

**Supplementary Figure 3.** Bar charts showing the percentage distribution of significant differentially methylated positions (DMPs) annotated to CpG island-related regions (A) and gene-associated regions (B) across each DNA methylation cluster. S_Shore: south shore; S_Shelf: south shelf; N_Shore: north shore; N_Shelf: north shelf; 3UTR: 3′ untranslated region; 5UTR: 5′ untranslated region; TSS200: 200 bp upstream of transcription start site; TSS1500: 1500 bp upstream of transcription start site.

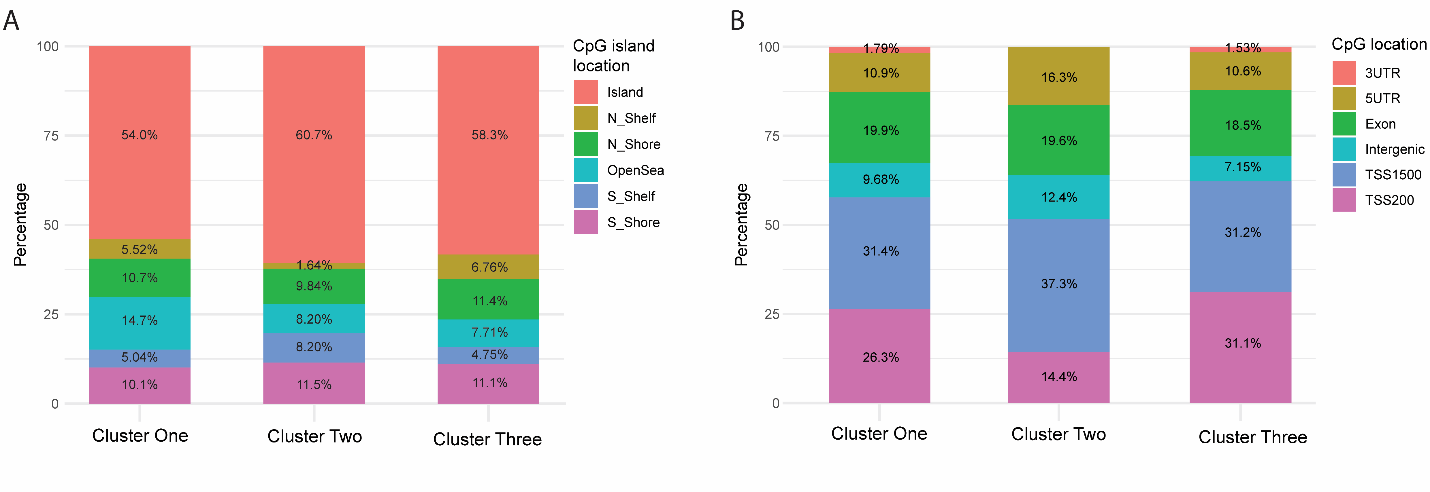
